# scRNAseq of thyroid eye disease orbital fat demonstrates fibroblast thyroid hormone signaling and SPARC production

**DOI:** 10.64898/2026.02.24.26346524

**Authors:** Emma JS Robinson, Kathrine Boest-Bjerg, Carlos Cuadros Sánchez, Sylvie Agnello, Alexandros Delimichalis, Gina-Eva Göertz, Insa Nolte, James A Pearson, Robert Andrews, Ilaria Muller, Ebony Smith, Liz Palmer, Jadwiga Furmaniak, Marian Ludgate, Peter N Taylor, Anja Eckstein, Sarah J Richardson, Catherine Rennie, Daniel S Morris, Anjana Haridas, Vickie Lee, Colin M Dayan, Stephanie J Hanna

## Abstract

There is an unmet need to identify biomarkers of active thyroid eye disease (TED). scRNAseq revealed that orbital fibroblasts from orbital decompressions in people with TED express high levels of thyroid hormone receptors, growth factor receptors, including insulin-like growth factor 1 receptor (IGF1R), and extracellular matrix proteins including SPARC (osteonectin), whereas orbital fat endothelial cells expressed thyroid peroxidase (TPO).

SPARC was significantly raised in the serum of people with thyroid disease compared to healthy controls. Furthermore, those with moderate, severe and sight threatening TED had higher SPARC levels than those with thyroid disease but free of TED or mild TED. Free-triiodothyronine (FT3) levels were positively correlated with SPARC in moderate-sight threatening TED. SPARC and IGF1 were positively correlated across people with thyroid disease alone, as well as TED. Thyroid stimulating hormone (TSH) levels were negatively correlated with SPARC in moderate-sight threatening TED. When participants were followed longitudinally, SPARC decreased after the active phase of TED. At the protein level, immunohistochemistry indicated that SPARC was heterogeneously expressed by fibroblasts in both control and TED orbital fat.

SPARC is a key mediator of fibrosis and deposition of extracellular matrix and the correlation of SPARC serum levels to TED status and FT3 make it a promising biomarker of active TED.

## Introduction

A subset of people with Graves’ disease (GD) develop thyroid eye disease (TED), where orbital inflammation coupled with overproduction of extracellular matrix (ECM) and expansion of adipose tissue result in the protrusion of the eyes, disfigurement and even blindness (1, 2). Antibodies that trigger GD are thought to cause TED, by binding thyrotropin receptor (TSHR) expressed in orbital fat (1). The beneficial effects of TSHR blockade on TED support this hypothesis (3). Signalling through the insulin-like growth factor 1 (IGF-1) receptor (IGF-1R) also contributes to TED and Teprotumumab which blocks activation of the IGF-1R, decreases the synergistic effect of IGF-1 signalling on TSHR signalling (4) and reduces the severity of TED (5).

A greater understanding of the pathological process is essential for novel treatments and early detection as TED is associated with substantial impairment of quality of life (6) and rapid diagnosis can enable earlier treatment opportunities which may be more effective especially when the disease is active.

However, for a disease found predominantly in those with hyperthyroid Graves’ disease (2, 7), there remains a paucity of information about thyroid hormones that can drive pathological processes in the orbital fat fibroblasts and the secretion of pro-inflammatory mediators by the fibroblasts. Furthermore, there remains an unmet need to identify biomarkers that assess the activity of TED. We profiled cells obtained from orbital decompressions in people with TED, using scRNAseq, and examined secreted factors in the serum, in order to address these questions.

## Results

### scRNAseq of orbital fat reveals a highly complex network of fibroblasts and extensive immune cell infiltration including plasma cells

To understand the functions of the fibroblasts in the orbital fat in TED and how they contribute to the autoimmune process, we digested orbital fat from TED decompression samples to single cells (n=4) and performed scRNAseq (Table 1). Large immune cell infiltrations, particularly plasma cells and B cells were noted as well as fibroblast and endothelial cell clusters (Fig 1A, Sup Fig 1). Major markers of the fibroblast subsets are shown in Fig 1B along with previously reported markers of fibroblasts in TED, including expression of TSHR at low level on lipo-fibroblasts. In particular we confirmed the presence of CD34+ fibroblasts that are progenitors for smooth muscle cells involved in connective tissue development and extracellular matrix binding (8, 9). Furthermore, a subset of FBLN1+ fibroblasts associated with connective tissue development were also present (8) as well as myofibroblasts. These are thought to differentiate from fibroblasts during TED and express proteins that can drive autoimmunity including IGF-1R (10, 11).

**Fig 1.**
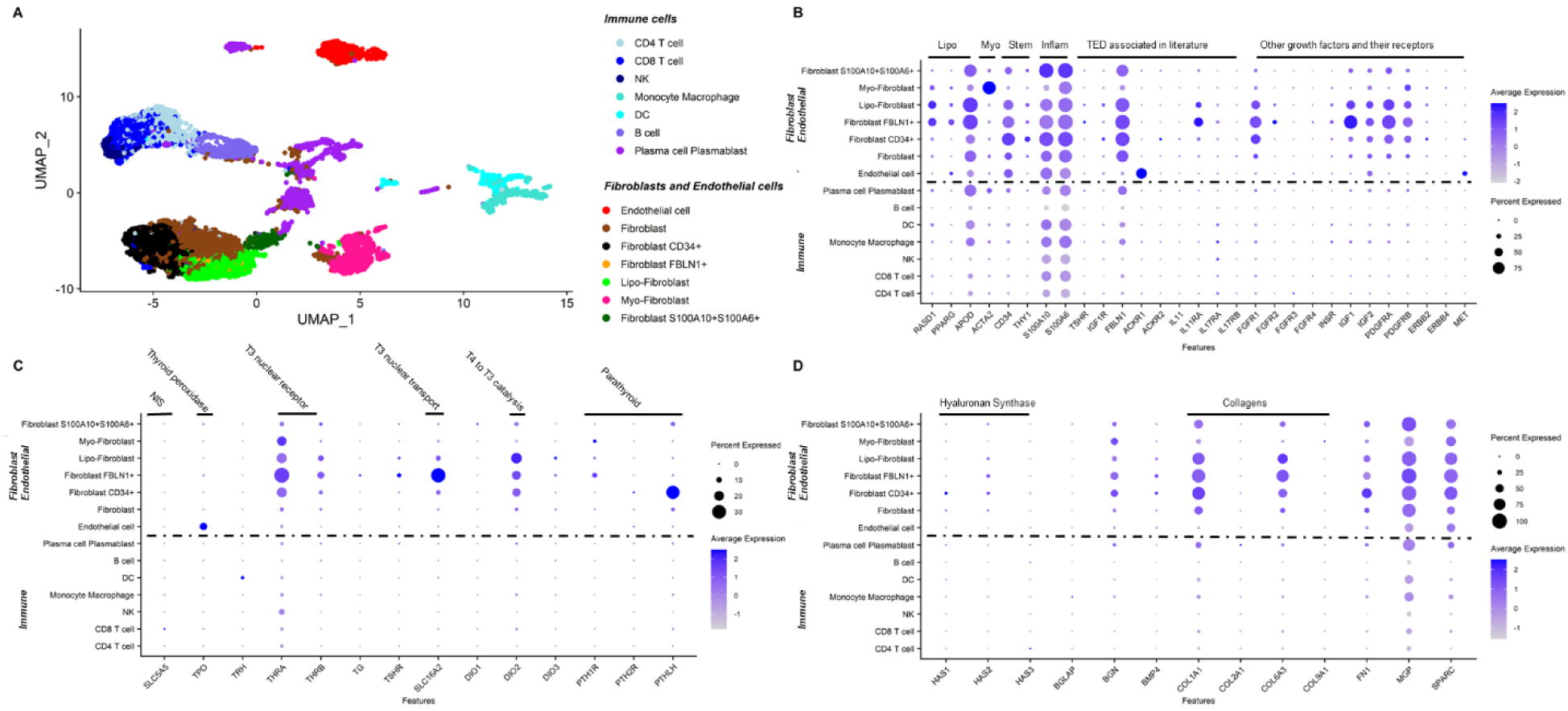
Orbital fat in TED contains many fibroblast subsets with distinct phenotypes, sensitivity to thyroid hormones and patterns of extracellular matrix production. A. UMAP plots of scRNAseq of orbital fat from TED. Cell identities were assigned using Azimuth, the Human primary cell atlas, the Fibroblast cell atlas and manual assignment and concatenated for further analysis. Data are combined from four individuals B. DotPlot of Major fibroblast subset markers, markers associated with TED in the literature and other growth factors and their receptors (in addition to IGF-1), coloured by average expression and scaled by percentage expressed. C. Thyroid and parathyroid hormone signalling and bone remodelling markers in orbital fat in TED. DotPlot of indicated gene expression in cells from orbital decompression, coloured by average expression and scaled by percentage expressed. SLC5A5: sodium/iodide symporter (NIS) TPO: Thyroid peroxidase TRH: Thyrotropin-releasing hormone THRA: Thyroid Hormone Receptor Alpha/Beta (nuclear receptor for T3) TG: thyroglobulin (T4 precursor) TSHR: TSH receptor SLC16A2: T4/T3 transporter. DIO1/2/3: dio-iodothyronine deiodinase 1/2/3 PTHLH: parathyroid hormone like hormone PTH1R: parathyroid hormone 1 receptor PTH2R: parathyroid hormone 2 receptor. (Not expressed: TSHB: TSH subunit beta, CGA: shared alpha subunit of TSH, PTH: parathyroid hormone) D. DotPlot of distinct pattern of production of extracellular matrix by orbital fat fibroblasts, coloured by average expression and scaled by percentage expressed.

**Table 1:** Characteristics and demographics of orbital fat donors.

| ID | Type | Age Bracket | Sex | Detail |
| --- | --- | --- | --- | --- |
| SC-A | TED | 50-59 | F | Decompression |
| SC-B | TED | 60-69 | F | Decompression |
| SC-C | TED | 50-59 | M | Decompression |
| SC-D | TED | 70+ | F | Decompression |
| CF-A | Control | 60-69 | M | Ptosis surgery |
| CF-B | Control | 60-69 | F | Blepharoplasty |
| CF-C | Control | 70+ | M | Enucleation |
| CF-D | TED | 50-59 | M | Blepharoplasty and fat pad debulking |
| CF-E | TED | 60-69 | M | Decompression for dysthyroid optic neuropathy |
| CF-F | TED | 50-59 | F | Decompression |
| CF-G | TED | 50-59 | M | Decompression for dysthyroid optic neuropathy |

### Gene expression in TED orbital fibroblasts indicates sensitivity to thyroid and parathyroid hormone signalling and bone remodelling

We wished to understand how thyroid and parathyroid hormone signalling influenced cells in the orbital fat as previous reports in the literature had indicated the ability of orbital fibroblasts to differentiate towards osteoblast-like phenotypes (12). We found that fibroblasts were enriched for THRA and THRB, the nuclear receptors for T3, SLC16A2 (the T4/T3 transporter) and DIO2 which catalyses conversion of T4 to T3, indicating that these cells are particularly sensitive to circulating levels of T4 (Fig 1C). Interestingly, at the mRNA level, relatively few fibroblasts expressed the TSHR, possibly indicating its stability at the protein level. Some fibroblast subsets expressed PTHR1, whilst others expressed PTHLH indicating a paracrine signalling process (Fig 1C, Sup Fig 1). In contrast other cell types such as B cells and T cells in the orbital fat showed no or very low level of expression of these genes. Notably, endothelial cells expressed high levels of TPO, the gene for thyroid peroxidase (for clarity, expression of THPO (thymopoietin) is also shown in Sup Fig 1).

### Fibroblasts are key producers of extracellular matrix in the orbital fat

Many *in vitro* assays study the ability of cultured orbital fat fibroblasts to produce hyaluronan. We leveraged the opportunity of the scRNAseq dataset and found that HAS2 (Hyaluronan Synthase 2) was widely expressed, with CD34+ fibroblasts also expressing HAS1. We also looked more broadly for extracellular matrix production and found a distinctive pattern of collagen, SPARC (osteonectin), BGN (biglycan), FN1 (fibronectin) and MGP (Matrix Gla Protein) expression (Fig 1D).

Because some of these proteins are also associated with bone remodelling, we next determined the expression of bone-remodelling markers. Analysis showed high expression of SPARC, COL1A1 and MGP, but lack of other canonical osteoblast markers such as ALPL (Sup Fig 2) in the fibroblasts.

**Fig 2.**
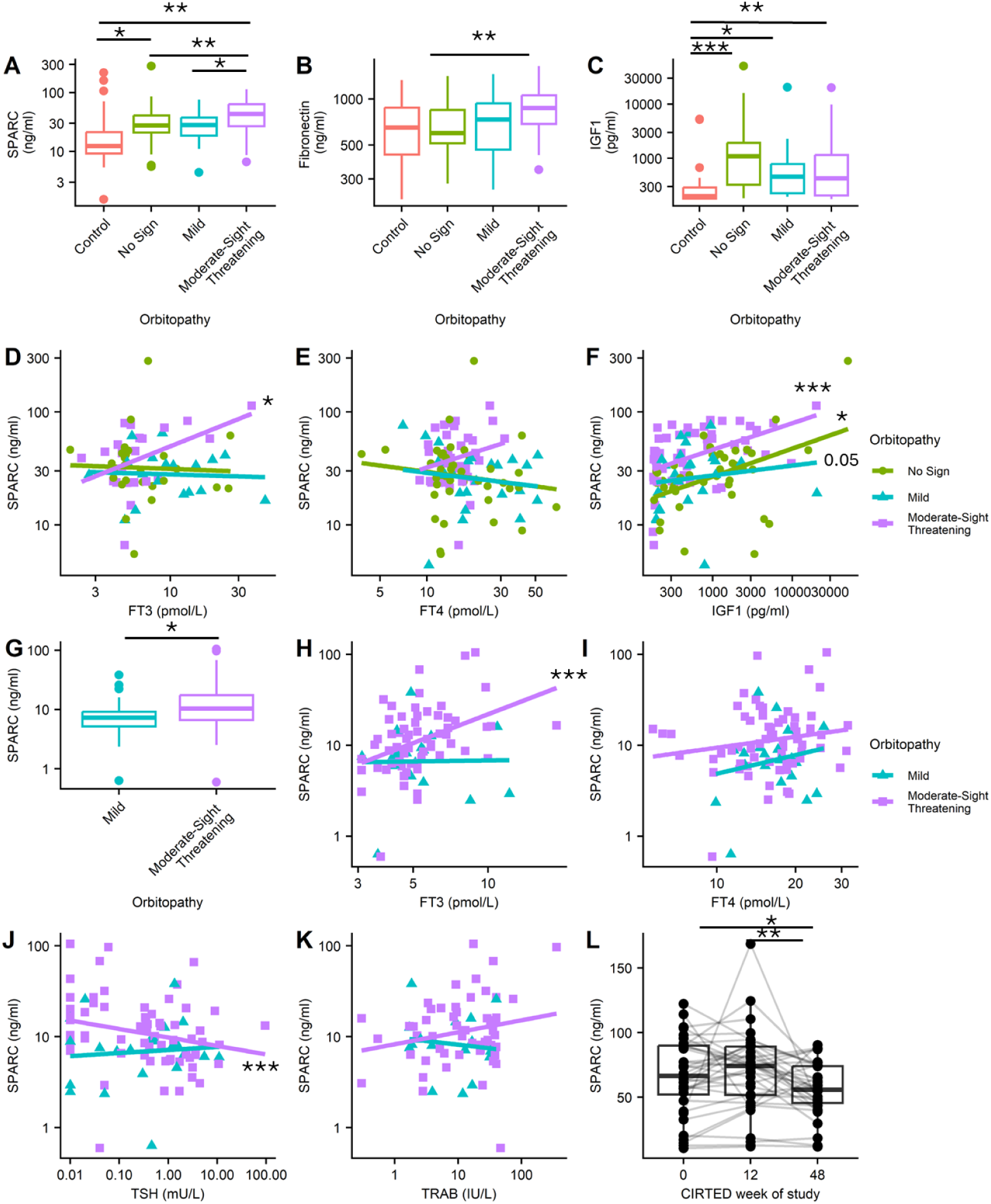
SPARC is raised in thyroid disease and further elevated in TED. Serum levels of fibroblast-secreted proteins. Split by orbitopathy level: A. SPARC B. Fibronectin, C. IGF1. Two tailed Wilcoxon signed rank test corrected for multiple observations were used to compare between all groups. SPARC correlations with D. FT3, E. FT4, F. IGF1 levels, coloured by cohort. Correlation by Spearman’s rank. G. SPARC in additional Essen cohort of serum. SPARC correlations with H. FT3, I. FT4, J. TSH, K. TRAB in Essen cohort. Two tailed T-test. L. SPARC over time in the CIRTED study. Two tailed Wilcoxon signed rank test corrected for multiple observations were used to compare between all groups. *p<0.05 **p<0.01 ***p<0.001

Recently, interest has focussed on targeting the neonatal Fc receptor in TED (13). We therefore looked for expression of Fc receptors in the orbital fat. Whereas resident immune cells expressed a heterogenous pattern of Fc receptors, fibroblasts expressed only large amounts of FCGRT, the neonatal Fc receptor (Sup Fig 3). It has also recently been noted that plasmablasts, plasma cells and B cells are present in TED (14). We examined the profile of antibodies expressed by fat-resident B cells, plasmablasts and plasma cells and found them to express all isotypes except IGHE, particularly IGHA1, IGHM and IGHG1 along with JCHAIN (Sup Fig 3).

**Fig 3.**
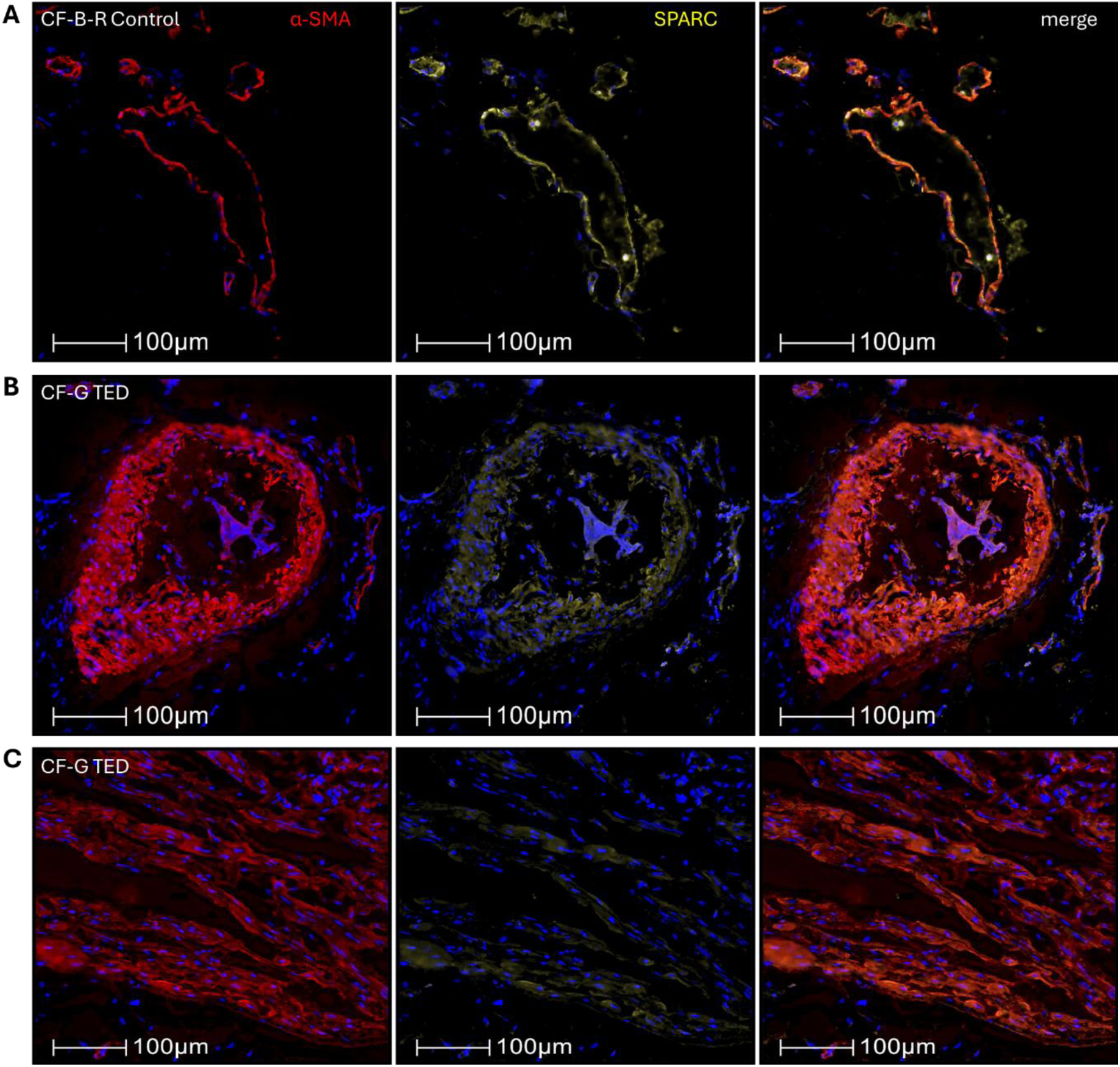
Heterogenous expression patterns of α-SMA and SPARC in orbital fat from control and TED donors. α-SMA (red), SPARC (yellow) and nuclei (blue) were stained in tissue sections and imaged. A. Control blood vessel and fibroblast locations of SPARC. B. TED blood vessel and fibroblast locations of SPARC. C. Dense tissue location of SPARC in TED sample. Images are representative of slides from three control donors and four donors with TED.

### Unique features of uncultured orbital fat fibroblasts in TED

It is understandably difficult to obtain control retro-orbital fat samples from healthy individuals and in addition, much work in the TED field has focussed on understanding fibroblasts that are expanded *ex vivo* and then differentiated towards adipocytes. To put our findings in context we compared our fibroblasts to those from a published dataset where orbital fat fibroblasts from TED were cultured for several passages and once they had sufficiently expanded (designated day 0 by the authors) they were differentiated to adipocytes (15). In addition, due to the high expression of CD34 in our fibroblasts, we utilised a published dataset of mesenchymal progenitors of adipocytes from white fat (16). Comparing all of the markers above, it was striking that the expression of thyroid hormone signalling pathway genes was particularly enriched in our freshly isolated TED fibroblasts, whereas for example FCGRT expression was high across all fibroblasts (Sup Fig 4).

### Serum SPARC is raised in people with thyroid disease and further elevated in people with TED

We hypothesised that signalling pathways in TED might include mediators from the fibroblasts being released into the blood. We noted Fibronectin (FN1) and SPARC as highly produced by orbital fibroblasts in the scRNAseq data. Serum analysis of these at the protein level identified that SPARC was significantly raised in people with thyroid disease in the CIRTED (48 participants), INDIGO (39 participants) and IDG (31 participants) cohorts of people with thyroid disease and thyroid eye disease (described in detail in Methods) compared to healthy controls (20 participants), (Fig 2A)(17, 18). Furthermore, those with moderate-severe or sight threatening (hereafter denoted moderate-sight threatening) TED had higher SPARC levels than those with thyroid disease but not TED or mild TED. Whilst serum Fibronectin showed similar trends, this was driven by significantly higher Fibronectin in the CIRTED cohort, with IDG and INDIGO not different to control (Fig 2B, Sup Fig 5).

### IGF-1 is raised in the serum of people with TED

We looked for mediators in the peripheral blood that orbital fibroblasts would be particularly sensitive to, based on our scRNAseq analysis, or those that have been previously reported to be elevated, namely IGF-1, PDGF-BB, PDL1, FGFbasic, FGF and FGF23 (19). We compared serum from healthy donors to donors with TED and those with thyroid disease but not TED from the three clinical studies of thyroid eye disease (CIRTED, IDG, INDIGO. We found that IGF-1 was raised in the serum of people with thyroid disease but not further raised in those with TED (Fig 2C). PDGF-BB was highest in those with thyroid disease in the INDIGO and IDG datasets but not CIRTED, and was not further raised in those with TED. None of the other three fibroblast growth factors nor PDL1 were consistently elevated in the groups with thyroid disease or TED (Sup Fig 6).

### Serum SPARC correlated with T3 and IGF-1

Interestingly, FT3 levels were positively correlated with SPARC in moderate-sight threatening TED (correlation co-efficient 0.430, p<0.05), although FT4 correlation did not reach statistical significance (Fig 2D,E). As SPARC is also reported to be secreted in response to IGF1 we also assessed correlation between these in the serum. SPARC and IGF1 were positively correlated across people with no TED (correlation co-efficient 0.39 p<0.05), mild TED (correlation co-efficient 0.39, p=0.05) and moderate-sight threatening TED (correlation co-efficient 0.5, p<0.01) (Fig 2F).

As we had examined multiple analytes without statistical correction, we validated our findings in a fourth dataset of observational serum samples of people with TED from Essen (109 participants). In these, again there was a significant increase in SPARC levels in people with moderate-sight threatening TED compared to those with mild TED (Fig 2G) (p<0.05). Furthermore, there remained a positive correlation with FT3 (Fig 2H) (correlation co-efficient 0.41 p<0.001), but not FT4 levels (Fig 2I) in moderate-sight threatening TED. In this cohort we also examined correlations with TSH, which exhibited a negative correlation with SPARC in moderate-sight threatening TED (correlation co-efficient -0.33 p<0.001), but not mild TED (Fig 2J). Thyroid stimulating receptor antibodies (TRAB) levels were not correlated with SPARC levels (Fig 2K).

### SPARC decreases over time in TED

We hypothesised that elevated SPARC in the serum was produced during the active phase of TED as it was highest in those with moderate-sight threatening TED and particularly high in CIRTED where people had new onset TED. We therefore measured SPARC in the serum of people in the CIRTED study longitudinally and found that SPARC remained high at 12 weeks and then significantly decreased at week 48 of the study (Fig 2L). This decrease occurred regardless of the treatment group (Sup Fig 7) and by week 48, CAS had decreased substantially for the majority of participants and no longer correlated positively with serum SPARC (data not shown). Four participants had serum from week minus 2 (prior to oral steroids) and baseline (week 0) and in these samples there was a trend towards decreased serum SPARC with steroid treatment (data not shown).

### SPARC distribution within orbital fat

We took further orbital fat samples from people with TED, both retroorbital and periorbital, and compared to control samples from people undergoing oculoplastic surgery for other indications (Table 1, Sup Fig 8). We imaged fixed sections of fat to examine spatial distribution of α-SMA as a marker of myofibroblasts and SPARC. It was notable in photos of the samples that TED fat is highly variable in appearance, with some samples appearing highly vascularised whereas others appeared more similar to white fat, even when samples were taken from both eyes of the same patient with bilateral TED (Sup Fig 8). This extended to imaging of the sectioned fat where distribution of cells, α-SMA and SPARC were highly heterogenous (Fig 3, Sup Fig 9). In both control and TED samples α-SMA and SPARC appeared in cells around blood vessels. SPARC was also present in isolated cells. Some TED samples had areas of dense fibrotic tissue which co-expressed SPARC and α-SMA. Although quantification of secreted, extracellular SPARC was not possible in these sections, this validated the expression of SPARC at the protein level in orbital fat fibroblasts in TED.

## Discussion

Our work highlights the importance of thyroid hormones and circulating growth factors in driving TED and confirms that pathogenic processes in orbital fat are highly complex. These involve not just TSHR or IGF1R signalling (as targeted by teprotumumab)(5) or chemokine and cytokine signalling, but T3, PTHLH and PDGF signalling, and result in elevated secretion of SPARC during the active phase of the disease.

It has previously been reported that fibroblasts from TED display increased features of osteogenesis upon ex-vivo differentiation including raised SPARC expression compared to control orbital fibroblasts (20). Our data demonstrated that SPARC was increased in the serum of people with thyroid disease and further raised in those with TED and this was consistent across the different cohorts. SPARC is also produced in bone by chondrocytes and osteoblasts where its production is significantly enhanced by T3 and IGF-1 (21, 22). We demonstrated that serum SPARC levels were significantly correlated with T3 levels only in people with moderate-sight threatening TED whereas SPARC levels were correlated with IGF-1 regardless of TED status. This suggests that the orbital fibroblasts in some patients with TED may have particular sensitivity to T3 levels, and suggesting an additional mechanism whereby achieving euthyroidism can prevent progression of TED(23). SPARC decreased by 48 weeks in the CIRTED cohort followed longitudinally, alongside an improvement in CAS scores. Imaging of control and TED orbital fat demonstrated highly heterogenous patterns of SPARC at the protein levels in different fibroblast subsets.

Li *et al.* recently published a scRNAseq analysis of orbital fat in TED. In a strong replication of their findings, we observed high expression of genes involved in growth factor signalling, including IGF1, FGF, EGF and PDGF. Li *et al.* also identified a large number of pathways associated with extracellular matrix formation, which again was replicated in our results and echoes findings in bulk RNA-seq (24, 25).

It is notable that we saw very few fibroblasts expressing TSHR, given that its expression in orbital fat is thought to drive TED and adipocyte differentiation(26) (27, 28) (29, 30). Due to our digestion and scRNAseq procedures we did not capture mature adipocytes(31) (32). Nonetheless it is surprising that TSHR expression was not more prominent. However, this replicates findings from a snRNAseq study where TSHR+ fibroblasts were relatively infrequent and not enriched in TED (33). This may be due to a stable expression at the protein level resulting in a low mRNA turnover(34). Furthermore, TSHR was enriched in fibroblasts with an FBLN+ or lipo-fibroblast phenotype (expressing the highest levels of PPARG). PTHLH was produced by fibroblasts whilst orbital fibroblasts also expressed PTHR1, indicating that some fibroblasts were programmed towards fibrosis rather than adipogenesis (35). High levels of T4 transporter/DIO2/THR expression found in our study indicate that the orbital fibroblasts will be particularly sensitive to circulating levels of thyroid hormones, underscoring the need to achieve euthyroidism to prevent development or progression of TED (23).

Except for TSHR we did not find expression of other thyroid specific proteins (thyroglobulin, thyroid peroxidase (TPO) or sodium-iodide symporter (NIS)) in orbital fibroblasts in this study. Expression of these four proteins has been well documented in fibrocytes/fibroblasts derived from healthy or patient donors (36, 37). The reasons why thyroglobulin, TPO or NIS were absent could be related to the low mRNA turnover for stably expressed proteins mentioned above. Expression of TPO by endothelial cells in the orbital fat was an unexpected finding and needs to be confirmed in further studies. However, this is a potentially relevant finding as interactions between TPO in the orbital tissue and anti-TPO antibodies may contribute to the pathogenesis of TED. Furthermore, anti-TPO antibodies could be useful markers of TED in TRAB negative disease (38, 39) (40)

We noted upregulation of many bone remodelling pathways in fibroblasts in TED, yet when we looked at key markers of chondrocytes, osteocytes, osteoblasts and osteoclasts, we saw a mixed picture across the fibroblast subsets, with COL2A1, biglycan, fibronectin, SPARC, COL1A1, MGP and CTSK particularly highly expressed. This is in agreement with reported expression of fibronectin and collagen by immunohistochemistry in orbital tissue samples

(41). However, others such as ALPL, a master regulator of osteoblast differentiation (42) were not highly expressed. Therefore, it would not be accurate to describe these fibroblasts as osteoblast-like, yet it is known that bony remodelling occurs in TED (43, 44) and these fibroblasts are likely contributing to this process. This also supports the theory that active TED is driven by stem cells. Expression of CD34 was high in many of the fibroblast clusters that we observed, suggesting overlap with previously reported phenotypes of infiltrating fibrocytes or mesenchymal stem cells (12, 45).

We considered the mechanisms that autoantibodies and T3 may use to drive remodelling processes. T3 stimulates osteoblast differentiation and bone formation and the production of SPARC (21, 46). In addition osteoblasts express TSHR and PTHLH (47, 48) and TSH promotes osteoblast proliferation and differentiation (49–52) (and causes upregulation of IL11R in osteoblasts) (53). Furthermore it has previously been shown that adipose-derived mesenchymal stem cells from the orbit have higher potential towards adipogenic and osteogenic differentiation but lower tendency to chondrogenesis when compared with abdominal ASCs (54, 55). In this context it is also interesting to consider the fact that thyroid nodules involve much extracellular matrix and ectopic calcium deposition (56).

This tendency towards secretion of bone remodelling proteins by orbital fibroblasts was particularly interesting in the context of smoking as a major risk factor for development of TED (1) and previous research demonstrating that smoking modulates expression of many bone proteins (57). Furthermore, bisphosphonates for treatment of osteoporosis and cancer can cause ocular side effects including orbital inflammation (58) and case reports of exacerbation of TED (59).

Previously it has been shown that FGFbasic and PDGF-BB synergistically induce hyaluron production and additively induce adipogenesis by orbital fibroblasts(60, 61). This is consistent with our scRNAseq data showing high levels of receptor for these growth factors on the orbital fibroblasts. Of the two, however, only PDGF-BB was elevated in the serum of people with thyroid disease in our study and was not further raised in TED. PDGFRA is raised in TED fibroblasts (62) and is particularly important in driving fibroblast-mediated connective tissue remodelling (63) and PDGFR signalling in osteoblast lineage cells controls bone resorption(64), linking the raised levels of PDGF-BB to raised levels of SPARC (and Fibronectin in the CIRTED cohort) that we also found in the serum of people with TED. This adds to a growing body of evidence that multiple inflammatory mediators and extracellular matrix proteins may be elevated in the blood in TED (25).

It has recently been demonstrated that patients with TED had higher levels of plasma FGF23 and sPD-L1 than those with Graves’ disease alone (19) however we were unable to replicate this in our current study. IGF-1 was also elevated in people with thyroid disease, highlighting the key role of the IGF1R in TED and emphasising the rational of targeting IGF1R with monoclonal antibodies. It should be noted that this was a simplified measurement as IGF-1 levels are affected by time of day and binding proteins (65). This evidence indicates multiple inflammatory programmes are occurring in TED, although IGF-1 signalling is thought to be upregulated early in orbital adipogenesis (15). Our research also reinforces a role for PDGF signalling as a possible target (66) in TED.

In summary, SPARC is a key mediator of fibrosis and deposition of extracellular matrix and here we show that it is highly expressed at both RNA and protein level by orbital fat fibroblasts in TED. A strength of this study was our demonstration that SPARC serum levels correlated to TED status across multiple independent clinical cohorts, making it a promising biomarker. Furthermore, for the first time this study suggests relationship between serum T3 levels and the mediator of fibrosis supporting the rationale for a strict control of thyroid function in the management of TED. Future work will aim to profile extracellular matrix proteins including SPARC, as well as growth factors and inflammatory mediators alongside FT3, FT4, TSH, TRABs (both stimulating and blocking) and αTPO antibodies, longitudinally in people with TED. Combined with detailed assessments of different components of clinical activity scores, with a focus on those indicating an active immune process, this will facilitate the prediction of disease trajectories and responses to therapies.

## Methods

### Sex as a biological variable

Our study examined males and females, and similar findings are reported for both sexes

### Participants-scRNAseq

Five patients with thyroid eye disease (TED) were recruited. Samples of peripheral blood, serum, and orbital fat were taken during surgery for planned or emergency orbital decompressions. One patient was excluded from scRNA-seq since they had a CAS of zero and few lymphocytes in preliminary flow cytometry analysis (data not shown). Serum taken on the day of decompression was analysed for FT3, TSH, FT4 and TSI (Thyroid stimulating immunoglobulin) at Cardiff and Vale University Health Board using standard procedures. Data about the samples can be found in Table 1.

### Participants- serum samples

Control serum was obtained from Cardiff University biobank from people with no history of autoimmune disease. Further serum was used from participants recruited under the IDG study ethics. The Immunological Determinants of Graves’ disease (IDG) study cohort included people with longstanding Graves’ disease but also lithium induced hyperthyroidism, toxic goitre, primary hyperthyroidism and Hashimoto’s thyroiditis. Of these a subset were recruited into the INDIGO study for those with either Graves’ disease- not on treatment, on treatment less than four weeks, new or active GO, recurrent Graves’ disease *(18)*. A further cohort was obtained from the Combined Immunosuppression and Radiotherapy in Thyroid Eye Disease (CIRTED) study which recruited people with active moderate-to-severe TED associated with proptosis or ocular motility restriction *(17)*. Serum and disease activity classifications from this cohort were used at baseline, week 0 (i.e. after two weeks of oral corticosteroids from week minus 2 to week 0, but before azathioprine/orbital radiotherapy treatment) for the serum ELISAs. Longitudinal serum samples were then taken at week 12 and week 48. For three participants where week 12 serum was missing, serum from week 14, 15 or 16 was used instead. Participants who had a baseline serum but were missing either week 12 or week 48 serum were included. In addition, participants who had a week 48 serum but were missing a week 48 CAS were included.

Demographics of the cohorts varied, as did the distribution of T3, T4 and TSH levels as set out in Table 2 and Sup Fig 10.

**Table 2:** Demographics of Control, CIRTED, INDIGO and IDG cohorts.

|  |  | TED Status |  |  |  |  |  |  |  |
| --- | --- | --- | --- | --- | --- | --- | --- | --- | --- |
| Cohort | n | Control | No Sign | Mild | Moderate-Sight Threatening | Female | Male | Age Median (years) | Age Range (years) |
| Control | 20 | 20 | 0 | 0 | 0 | 15 | 5 | 44.5 | 25-66 |
| CIRTED | 48 | 0 | 0 | 7 | 41 | 36 | 12 | 52.7 | 25.4-69.6 |
| IDG | 31 | 0 | 26 | 5 | 0 | 24 | 7 | 46.5 | 16.6-73.4 |
| INDIGO | 39 | 0 | 15 | 13 | 11 | 32 | 7 | 43.3 | 19.9-74.3 |

Serum samples from the Essen Cohort were obtained from the University Duisburg-Essen Biobank for patients with Graves’ disease and TED. Thyroid eye disease (TED) classification was evaluated using the NOSPECS and stratified as mild (NOSPECS <5) or moderate-severe (NOSPECS ≥5). Clinical assessments were performed by an ophthalmologist with expertise in TED.

One profoundly hypothyroid participant from Essen was included in the initial measurement of serum SPARC but excluded from correlation analysis.

### FT3, FT4 and TSH measurements

Measurements were used from the original datasets (17, 18). FT4, FT3 and TSH in the IDG study were measured by Abbott Alinity Immunoassay. Serum levels of FT4, FT3, and TSH in the Essen cohort were measured using the Siemens Atellica® IM Immunoassay. TRAb levels were measured using a third-generation Roche assay. TSI were measured using a Siemens Immulite Thyroid stimulating Immunoglobulins chemiluminescent immunoassay.

### Participants- Immunofluorescent imaging

Patients undergoing oculoplastic surgery for TED or non-TED indications were enrolled into the Immunological determinants of Graves’ disease study and discarded fat from their surgery was collected. Patient details and photos of fat obtained are shown in Table 1 and Sup Fig 8.

### Sample processing scRNAseq

To isolate single cells from the orbital fat, it was cut into small (2-3mm diameter) pieces and digested for 15 minutes in Hank’s buffered saline solution (HBSS) containing 2mg/ml collagenase P, 1mg/ml dispase and 1ul/ml DNaseI (100u/ml) at 37°C with shaking. Samples were centrifuged and the fraction containing PBMCs and fibroblasts was frozen in CryostorCS10 (StemCell Technologies). Samples were then thawed, washed (BioLegend) and run on a 10xgenomics Single Cell 5’ Reagent kit v2. Samples were sequenced on an Illumina NextSeq, using a target read depth of 20,000 reads/cell for GEX and 5,000 reads/cells for cell hashing.

### scRNAseq data analysis

CellRanger 6.1 (10X Genomics), was used to align the reads obtained from sequencing. The rest of the analysis was performed using R 4.1.1. The code was run on RStudio IDE version 2022.07.01, build 554. Seurat version 4.1.1 was used for the data processing with parameters FindVariableFeatures= 2000 nfeatures, KNN= 18 dimensions, FindClusters resolution=1.2(67). Doublets were removed by Seurat quality control and doubletFindeR version 2.0.3 (68). Using the PCA data, the batch effect was corrected using Harmony version 0.1.0 (69). K-nearest neighbours were performed using the FindNeighbours function for the 20 first dimensions from the harmony reduction. Finally, the data were clustered using the FindClusters function with resolution argument 0.5. PBMC subsets were identified using the Azimuth reference dataset and manual gating (67).

The Human Primary Cell Atlas (70) was used to identify other cell types in the data using Celldex version 1.4.0, using SingleR 1.81. Fibroblasts were identified using the Fibroblast Atlas (71) (GitHub repository (https://github.com/immunogenomics/fibroblastatlas and Symphony, https://github.com/immunogenomics/symphony)

To compare our freshly isolated samples to other fibroblasts, datasets from Kim *et al* (15) and Merrick *et al* (16) were accessed from NCBI Geo, processed using CellRanger 6.1 and integrated with Seurat and Harmony.

### Serum ELISAs

ELISAs were from Biolegend (FGF basic) and R&D/Biotechne (BMP4, IGF1, Fibronectin, PDL1, FGF7, FGF23, SPARC, PDGF-BB duosets). Serum was tested undiluted except for the longitudinal CIRTED, Essen and belimumab study samples which were diluted 1:2 in PBS. In all assays the median values fell within the standard curve range provided by the manufacturer. Standard curves were extrapolated to quantify absorbance values above the range. For PDL1 and FGF7, absorbances below 0.5 x the lowest standard curve value were assigned a value of 0.5 x the lowest standard curve concentration. Where there was insufficient serum to run all assays, serum was randomly assigned to assays.

### Orbital fat fixation

Orbital fat tissue was fixed in Periodate-lysine-paraformaldehyde (PLP) for 16-20 hours, prior to infusion with 10% sucrose, then 20% sucrose, for 30 mins each (all steps at 4°C). Tissues were subsequently blocked in optimal cutting temperature (OCT) media using an isopentane bath.

### Immunofluorescent labelling and imaging

Orbital fat tissues were embedded in optimal cutting temperature medium and sectioned at 5-um thickness. Sections were air dried and fixed for 30 minutes with 1% paraformaldehyde at room temperature. Following fixation, tissue was permeabilised with 0.1% Triton X-100 for 30 minutes and blocked for 1 hour with 5% BSA and 5% NGS, before overnight incubation at 4 °C with rabbit anti-human SPARC (HPA002989, Sigma) and mouse anti-human aSMA (ab7817, Abcam). Secondary labelling was performed for 1 hour with both Alexa Fluor 647-conjugated goat anti-rabbit antibody (A21245, Invitrogen) and Alexa Flour-555 goat anti-mouse antibody (A21424, Invitrogen) as well as DAPI. Slides were mounted with ProLong Diamond Antifade Mountant (P36961, Invitrogen). Slides were scanned at x40 magnification using the MOTiF workflow on a PhenoImager whole slide scanner (Akoya Biosciences).

### Statistics

Data were analysed in R version 4.4.2 using packages tidyverse, ggplot2, cowplot, scPubr, Seurat, DoubletFinder, readxl, haven, tidyr, ggpubr, celldex, SingleR, symphony, drc, stats, rstatix, dplyr.

ELISA standard curves were constructed using drc::drm and unknown values calculated using stats::predict

Differences between groups in serum ELISAs were calculated using rstatix in R with a two-sided wilcox_test and p.adjust.method holm for multiple comparisons. For the longitundinal study of CIRTED a one way ANOVA was performed.

Correlation tests were performed with cor_test from the rstatix package using method = "spearman", use= "pairwise.complete.obs" and for sub-analysis of individual cohorts with moderate to sight threatening TED only, alternative="g".

### Study approval

The Immunological Determinants of Graves’ Disease (IDG) study was approved by the South Wales Research Ethics Committee (REC reference: 12/WA/0285 Protocol number: SPON1106-12). INDIGO was a sub-study of IDG (Marie-Sklodowska Curie Industry Industry-Academia Pathways and Partnerships (IAPP) action, GA number 612116.

The Combined Immunosuppression and radiotherapy in thyroid eye disease (CIRTED study) was approved by the NHS South West Central Bristol Research Ethics Committee reference 05/Q2006/62, MHRA reference 03299/0003/001-0001, and local approval was obtained to use serum in the current study.

Healthy donor serum samples were provided through the Cardiff University Biobank (Application Number 24/0002).

Serum samples from the Essen Cohort were obtained from the University Duisburg-Essen Biobank for patients with Graves’ disease and TED in accordance with approval from the Institutional Ethics Committee. All samples were collected in accordance with approval from the Institutional Ethics Committee (06–3211 and 14–5965-BO).

All studies were conducted in accordance with the principles of good clinical practice established by the International Council for Harmonization/WHO. All participants provided written informed consent prior to enrolment, as mandated by the Declaration of Helsinki. This study was carried in accordance with principles of Good Clinical Practice and in accordance with all applicable regulatory requirements

## Supporting information

Raw Data

## Data Availability

scRNAseq data will be made available in a public repository at point of publication. All other data are available from the corresponding author upon reasonable request.

## Notes

All other authors declare no conflict of interest

### Competing Interest Statement

IM - Advisory board; Amgen & Merck, Consulting; Tourmaline VL- Advisory Board: Amgen & Viridian; Principal Investigator: Horizon, Sling, Lassen, Argenx & Viridian Trials. CMD- Advisory board/lecturing: Lundbeck, Immunovant, Lassen, Amgen, Argenx, Sling/Vasaragen. ML-Advisory board/lecturing Ethyreal, Aspen/Meridia, PNT Advisory board/lecturing: Immunovant, ECLOSIX, IBSA
All other authors declare no conflict of interest

### Funding Statement

This research was funded by Fight for Sight/ Thyroid Eye Disease Charitable Trust small grant RESSGA2310 and seedcorn funding from Cardiff University SIURI. SJH is funded by the Diabetes and Wellness foundation Professor David Matthews Fellowship. JAP and KBB are funded from a Medical Research Council Career Development Award to JAP (MR/T010525/1). SA is funded by a Medical Research Council Doctoral Training Partnership PhD Award.

### Author Declarations

The Immunological Determinants of Graves Disease (IDG) study was approved by the South Wales Research Ethics Committee (REC reference: 12/WA/0285 Protocol number: SPON1106 12). INDIGO was a substudy of IDG (Marie Sklodowska Curie Industry Industry Academia Pathways and Partnerships (IAPP) action GA number 612116. The Combined Immunosuppression and radiotherapy in thyroid eye disease (CIRTED study) was approved by the NHS South West Central Bristol Research Ethics Committee reference 05/Q2006/62 MHRA reference 03299/0003/001 0001 and local approval was obtained to use serum in the current study. Healthy donor serum samples were provided through the Cardiff University Biobank (Application Number 24/0002). Serum samples from the Essen Cohort were obtained from the University Duisburg Essen Biobank for patients with Graves disease and TED in accordance with approval from the Institutional Ethics Committee. All samples were collected in accordance with approval from the Institutional Ethics Committee (06 3211 and 14 5965 BO).

