## Supplemental Figures for "scRNAseq of thyroid eye disease orbital fat demonstrates fibroblast thyroid hormone signaling and SPARC production"

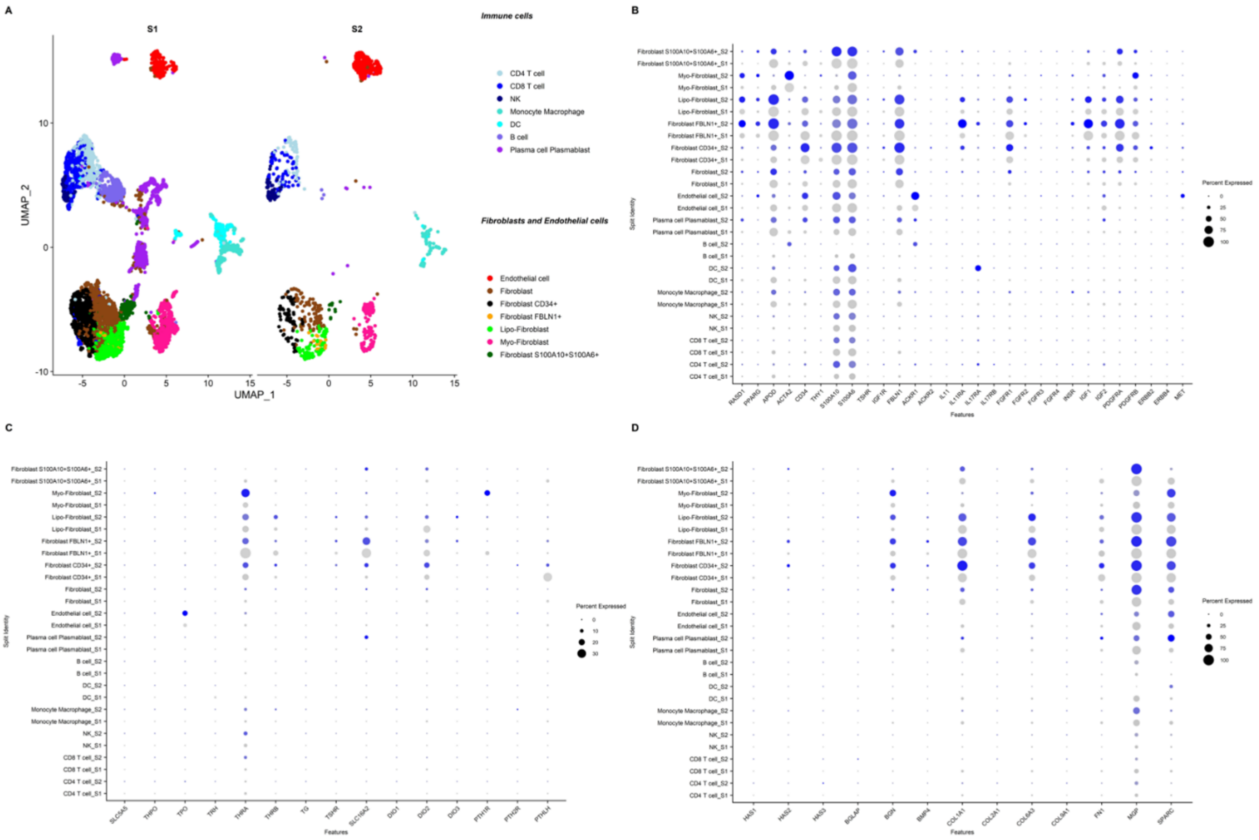


**Sup Fig 1. Orbital fat in TED contains many fibroblast subsets with distinct phenotypes, sensitivity to thyroid hormones and patterns of extracellular matrix production** A. UMAP plots of scRNAseq of orbital fat from TED. Data are split between two samples, each pooled from two donors. B. DotPlot of Major fibroblast subset markers, markers associated with TED in the literature and other growth factors and their receptors (in addition to IGF-1), scaled by percentage expressed and split by sample. C. Thyroid and parathyroid hormone signalling and bone remodelling markers in orbital fat in TED. DotPlot of indicated gene expression in cells from orbital decompression, scaled by percentage expressed and split by sample. D. DotPlot of distinct pattern of production of extracellular matrix by orbital fat fibroblasts, scaled by percentage expressed and split by sample. THPO (thrombopoietin) is also shown for clarity.

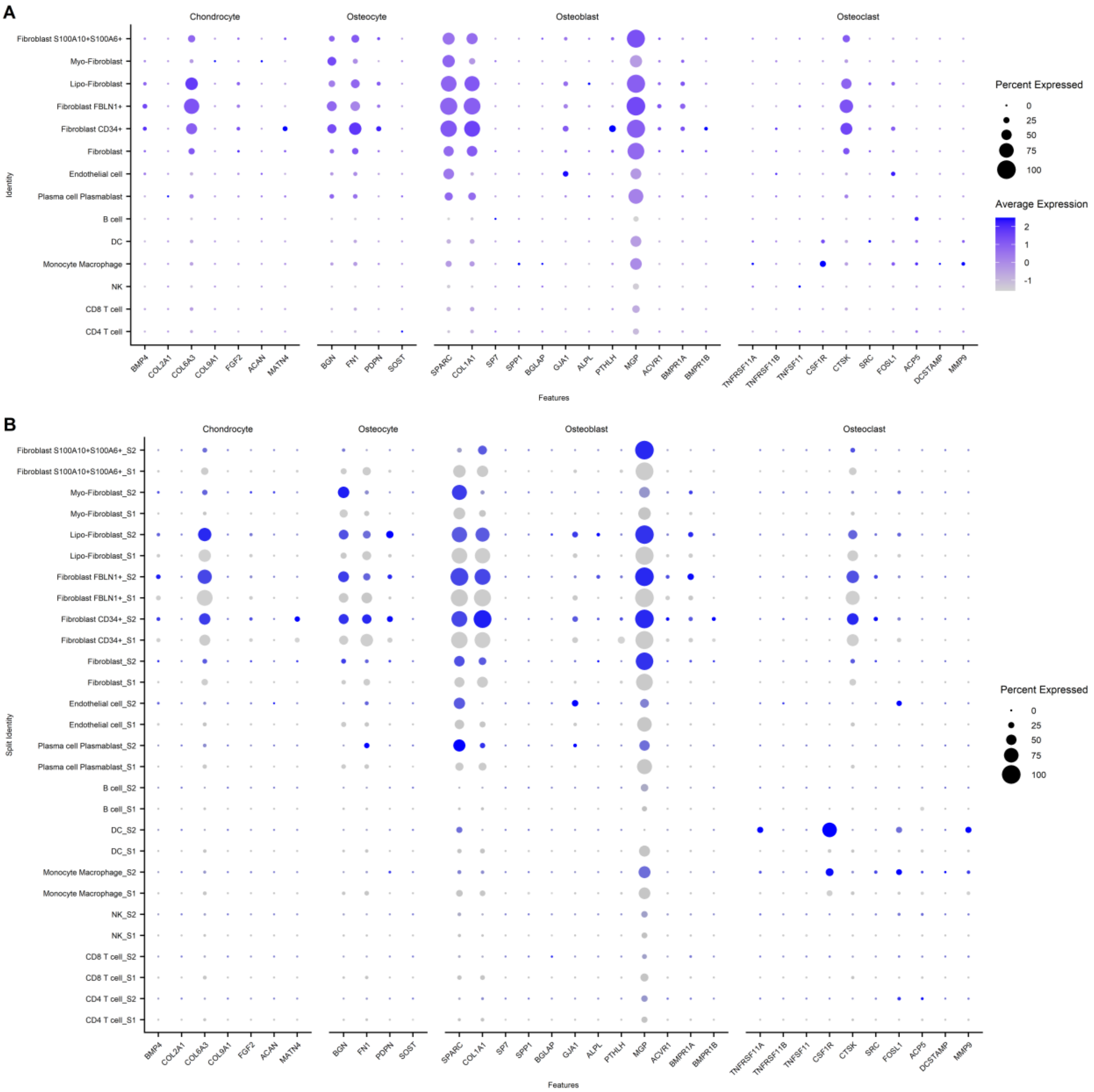


**Sup Fig 2. Orbital fibroblasts have features of tissue remodelling but do not express canonical osteoblast markers**. Dot plots of bone remodelling markers in orbital fat in TED scaled by percentage expressed and A. Coloured by expression level B. Split by sample

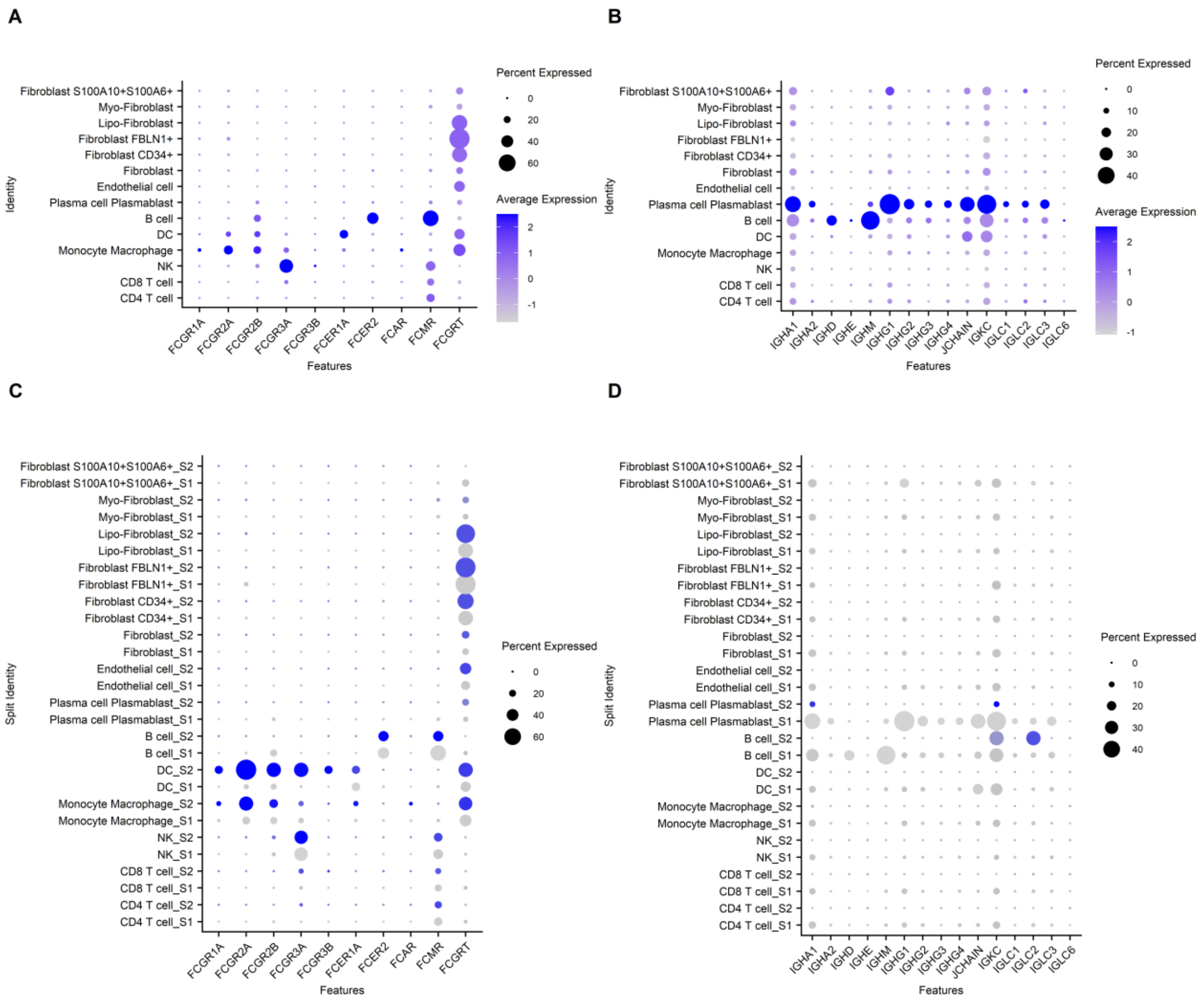


**Sup Fig 3 The neonatal Fc receptor is highly expressed in orbital fibroblasts in TED, whilst tissue-resident B cells, plasmablasts and plasma cells secrete antibodies.** A. DotPlot of Fc receptor expression in orbital fibroblasts and infiltrating immune cells, scaled by percentage expressed and coloured by expression level. B. DotPlot of immunoglobulin chain expression, scaled by percentage expressed and coloured by expression level. **C.**DotPlot of Fc receptor expression in orbital fibroblasts and infiltrating immune cells, scaled by percentage expressed and split by sample. B. DotPlot of immunoglobulin chain expression, scaled by percentage expressed and split by sample.

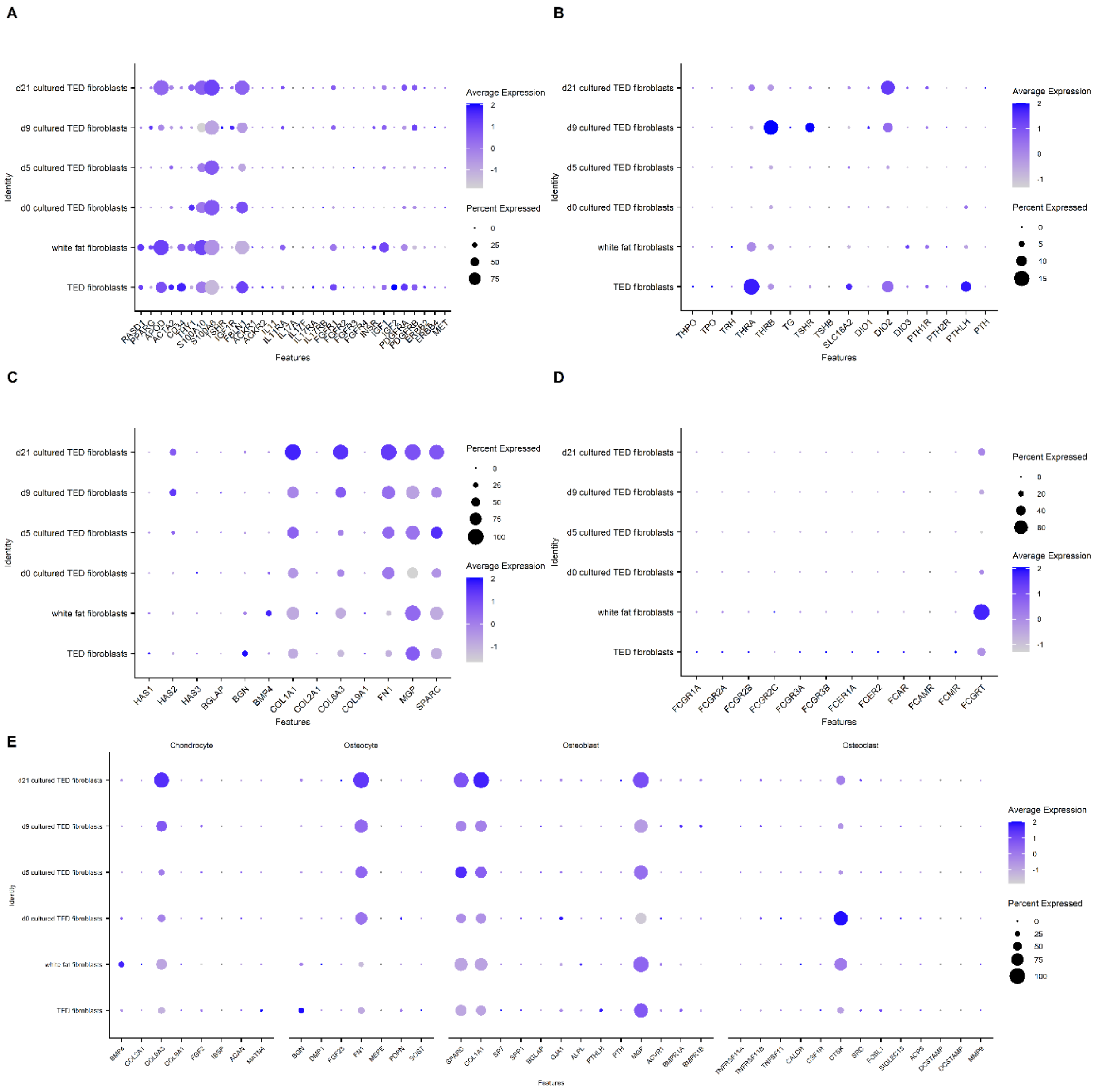


**Sup Fig 4: Unique features of orbital fat fibroblasts in TED.** scRNAseq of fibroblasts from the people (TED fibroblasts), were compared to scRNAseq of freshly isolated mesenchymal cells including those with preadipose, endothelial and smooth muscle phenotypes analogous to our fibroblast subsets from a publicly available dataset. In addition, they were compared to scRNAseq of fibroblasts cultured for several passages from TED orbital fat (d0 cultured TED fibroblasts) and then differentiated towards adipocytes for 5,9 or 21 days in a publicly available dataset. A. Canonical fibroblast markers. B. Thyroid hormone signalling C. Extracellular matrix production D. Fc receptors E. Bone morphogenic markers. Dotplots are coloured by expression level and sized by percentage expression.

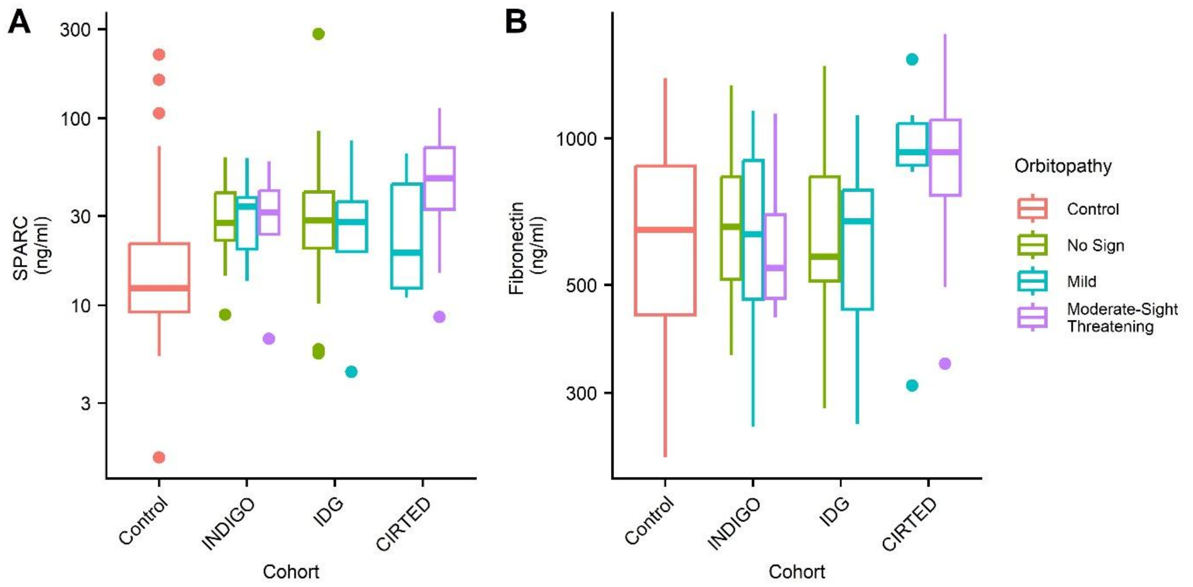


**Sup Fig 5. SPARC and Fibronectin levels by cohort** Serum levels of fibroblast-secreted proteins. Split by orbitopathy level and cohort: A. SPARC B. Fibronectin, corrected and uncorrected significance levels are in Sup Table 1.

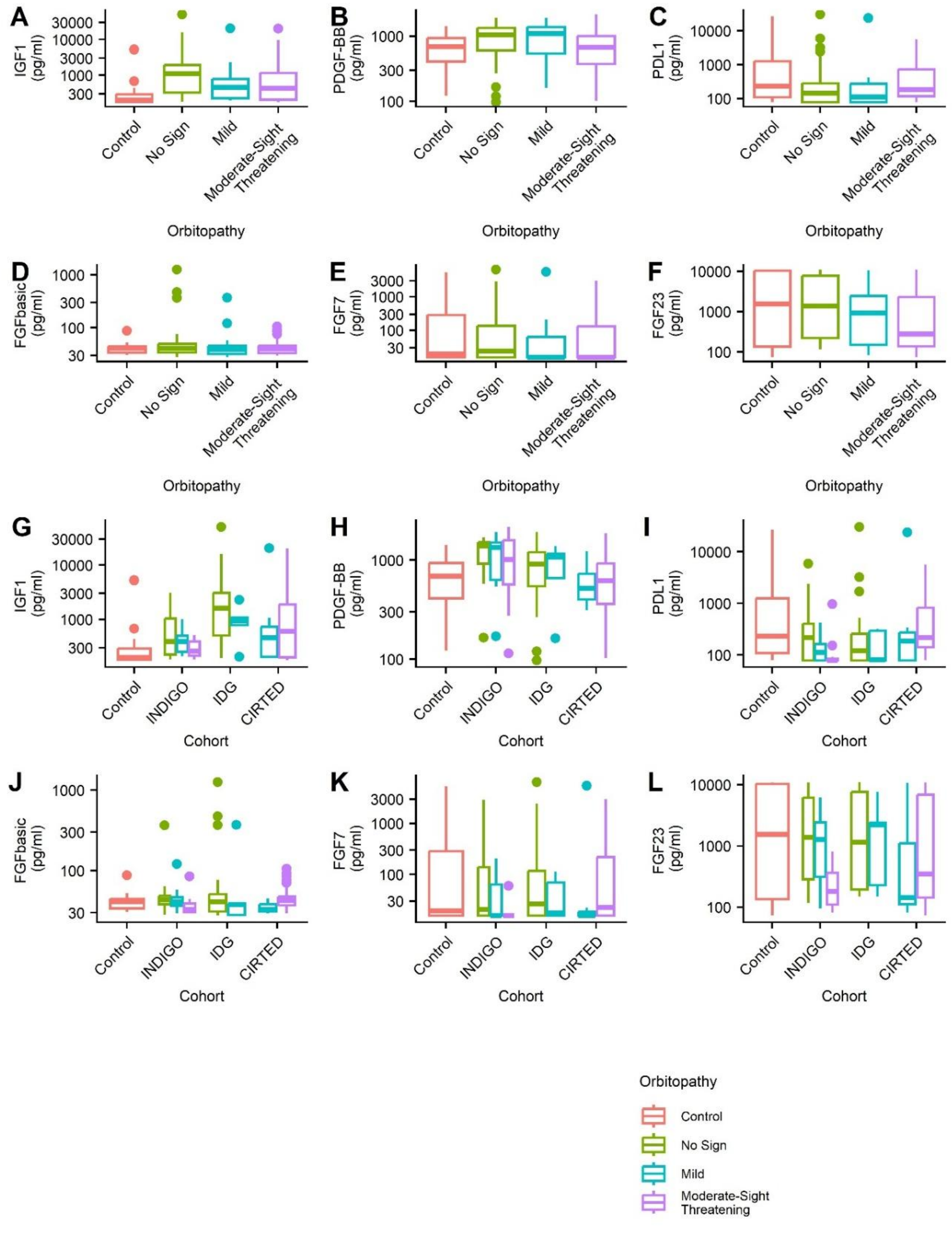


**Sup Fig 6. Growth Factors and PDL1 in the serum.** Split by orbitopathy level: A. IGF1 B. PDGF-BB, C. PDL1, D. FGFbasic, E. FGF F. FGF23 Two tailed Wilcoxon signed rank test corrected for multiple observations were used to compare between all groups. Split by orbitopathy level and cohort: G. IGF1 H. PDGF-BB, I. PDL1, J. FGFbasic, K. FGF L. FGF23 Corrected and uncorrected significance levels are in Sup Table 1

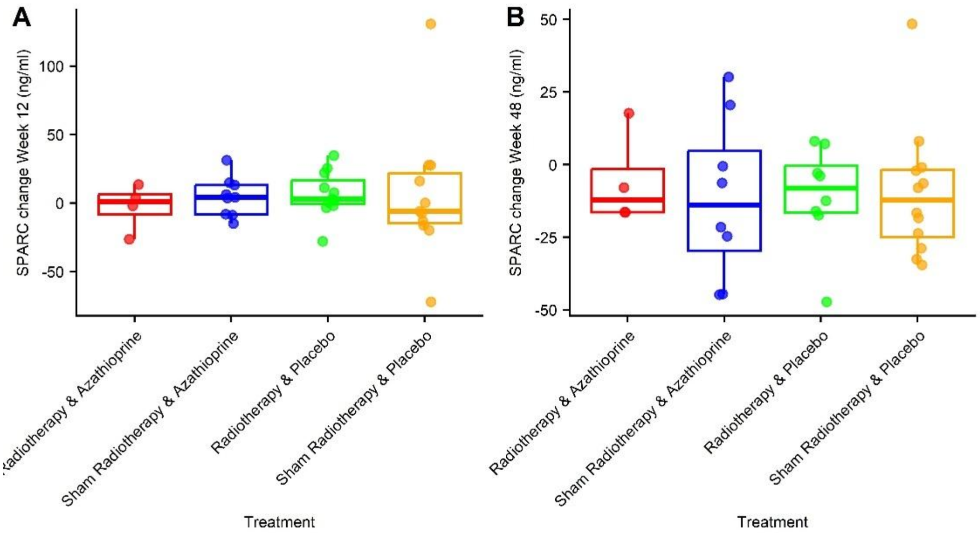


**Sup Fig 7. Timecourse of SPARC reduction was not affected by type of therapy.** The decrease in SPARC from week 0 to A. week 12 and B. week 48 is shown for individual participants in the CIRTED study, split by treatment group.

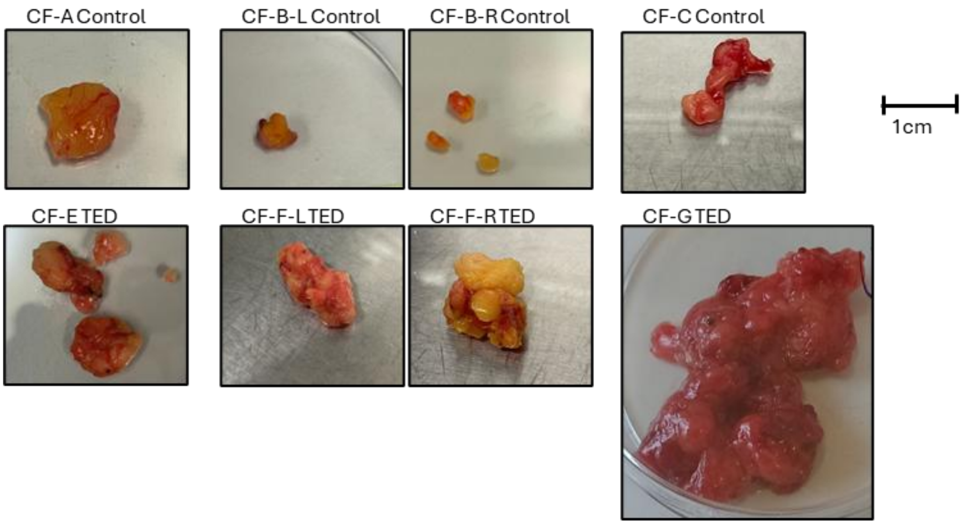


**Sup Fig 8. Photographs of orbital fat samples used for SPARC imaging.** The participant ID and left (L) or right (R) eye is indicated above each image. No image was available for CF-D. All photographs are shown to the same scale.

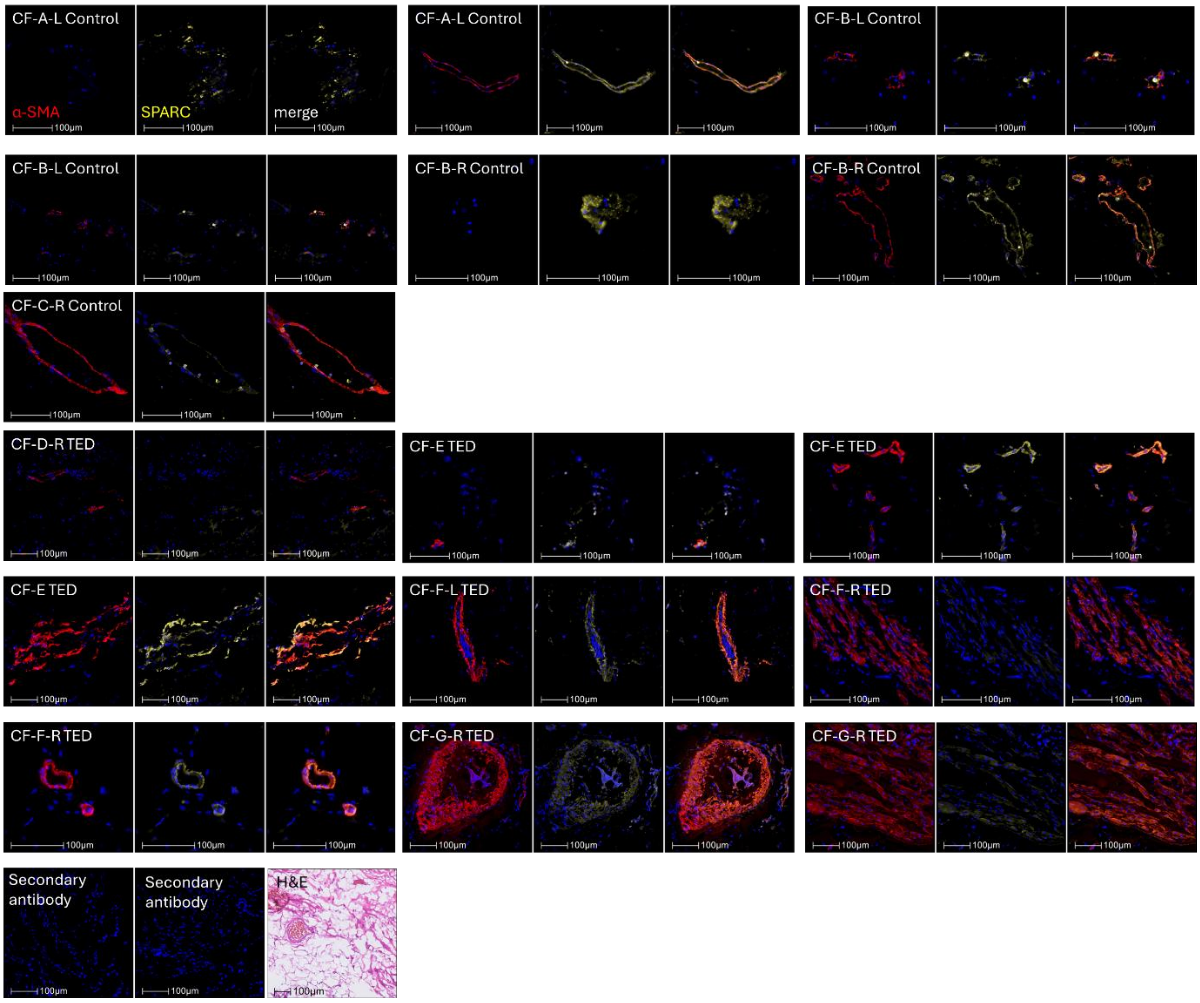


**Sup Fig 9. Heterogenous expression patterns of α-SMA and SPARC in orbital fat from control and TED donors.** α-SMA (red), SPARC (yellow) and nuclei (blue) were stained in tissue sections and imaged. Representative images of the heterogenous distribution and expression of SPARC are shown in each of the three control and four TED donors.

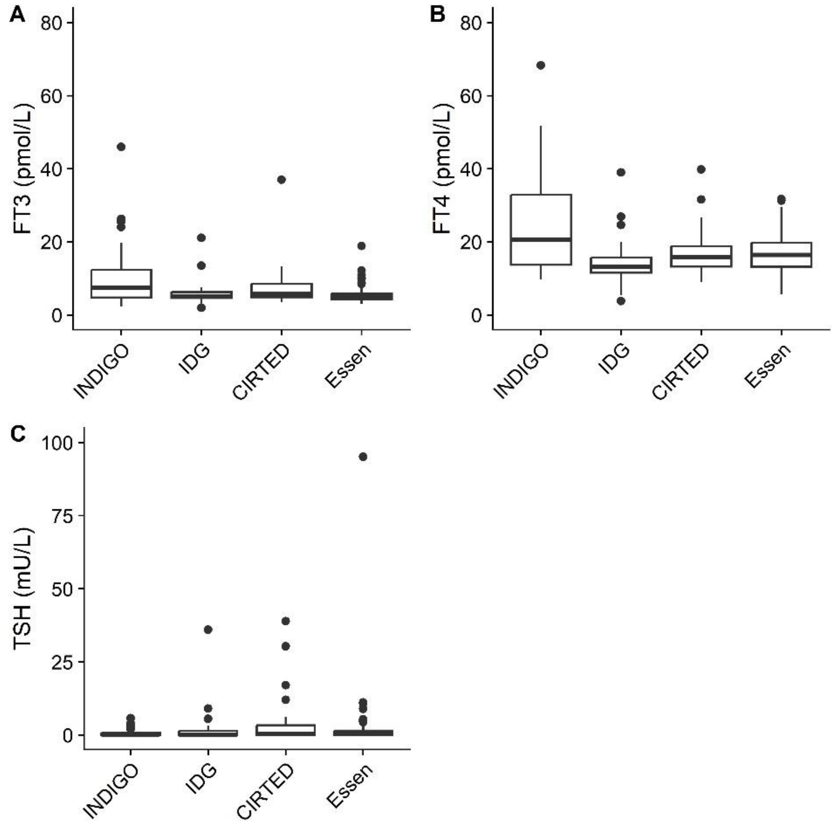


**Sup Fig 10. T3, T4 and TSH in the cohorts.** A. FT3, B. FT4, C. TSH. Reference ranges TSH 0.3-4.4mU/L, fT4 9-19.1 pmol/L, FT3; 2.4-6.0 pmol/L. The central line represents the median. The lower and upper hinges correspond to the first and third quartiles. The upper whisker extends from the hinge to the largest value no further than 1.5 * IQR from the hinge. The lower whisker extends from the hinge to the smallest value at most 1.5 * IQR of the hinge. Data beyond the end of the whiskers are plotted individually.
